# The tiered restrictions enforced in November 2020 did not impact the epidemiology of the second wave of COVID-19 in Italy

**DOI:** 10.1101/2021.09.02.21263035

**Authors:** Maurizio Rainisio

## Abstract

In the attempt to counter the second COVID-19 wave in autumn 2020, the Italian government introduced a unique countrywide system of physical distancing measures organized in progressively restrictive tiers (coded as yellow, orange, and red) imposed on a regional basis according to epidemiological risk assessments. This followed a set of other scattered or limited measures described in full detail by Manica et al.^1^.

This paper concerns the introduction of the tiered restrictions. The data used to evaluate the impact of the restriction system on SARS-CoV-2 transmissibility and hospitalization burden in Italy is from the Italian COVID-19 integrated surveillance system.

The reproduction number R(t) estimated on the onset of COViD-19 symptoms constantly decreased during most of the observation period 04 October 2020 - 06 December 2020 than is centered on the date of introduction of the measures and includes the expected latency to reach an effect. The fastest decrease of R(t) occurred in the period preceding the introduction of the tiered restrictions. There was no evidence that introducing the tiered restrictions determined the reduction of R(t). An alternative analysis using the daily growth rate of the epidemic curve confirms these conclusions. Similar considerations are made concerning the health system burden, analyzing hospitalizations at country level.

The trend of R(t) tending to increase shortly after the measures became effective does not allow to exclude that the enforcement of such restrictions might have been counterproductive.

These results are instrumental in informing public health efforts aimed at attempting to manage the epidemic efficiently. Planning further use of the tiered restrictions and the associated containment measures should be carefully and critically revised to avoid a useless burden to the population with no advantage for the containment of the epidemic or a possible worsening.

## Material

Istituto Superiore di Sanità (ISS) provides a daily updated Excel file^2^ reporting summary and raw data for a few parameters. Among the available raw data, the incidence of new COViD-19 cases (casi sintomatici) was analyzed as well as the number of hospitalizations. Unfortunately, the data provided to the public, including researchers not associated with the institute, is only pooled at the national level. The download of 31 August 2021 was used.

The daily data provided by Protezione Civile (PC)^3^ that includes data at the regional level for the incidence of new infections (nuovi_casi) from the download of 31 August 2021 was used.

While estimates are graphically reported only for the period 04 October 2020 to 06 December 2020, they were computed on the totality of the data. The reporting period was selected post hoc to include the whole period of decrease of R(t) for most of the cases that have been considered.

## Methods

As a measure of transmissibility, the net reproduction number R(t)^4^ is considered.

R*(t) was computed using the R program EpiEstim^5^ for three parameters: the daily number of new symptomatic cases and new hospitalizations from the ISS database, and the number of new detected SARS-CoV-2 infections from the database of PC. For the latter, in addition to the pooled data at the country level, the data of the 9 Italian regions with more than two million inhabitants covering (80.0% of the country’s population) were used.

Estimates of R^*^(t) were computed using the distribution of the serial interval estimated from the analysis of contact tracing data in Lombardy^6^ (a gamma function with shape 1.87 and rate 0.28, for a mean of 6.68 days) as a proxy for the distribution of the generation time.

Estimates of R^*^(t) are attributed to the start of the seven days interval used to perform the estimation; this allows to correctly attribute R^*^(t) = 1 to the maxima/minima of the epidemic curve I(t). The estimates of R^*^(t), while computed on 7-day intervals, are still affected by excessive random variability due to the imprecision of the original data. R^*^(t) has been approximated by a thin plate regression spline^7^ using SAS^®^-PROC TPSPLINE with smoothing factor log_10_(nλ) = 2; this value was chosen arbitrarily to adequately smooth the empirical curve. The resulting function R^S^(t) helps further study the R*(t) trend. The 1^st^ derivative of R^S^(t) allows the approximation of maxima and minima of R^*^(t) that identify the time when any intervention might have influenced the epidemic trend changing the sign of the curvature of I(t); inflection points identify the values of t when a local maximum acceleration for decrease or increase happened.

As a supportive analysis, the ratio between two consecutive days of the epidemic curve was considered. It matches the estimates of R*(t) qualitatively while using a conceptually more straightforward framework. I^S^(t) is defined as the thin plate spline approximation of I(t); log_10_(nλ) = 3 was empirically identified as the smallest smoothing factor able to eliminate the pronounced weekly seasonality of the data.

G^S^(t) = (I^S^(t)/I^S^(t-1))^7^ is the ratio under study. The power 7 was introduced to match R^*^(t) values and make the data easier to communicate as estimates of the weekly rates.

The derivative of G^S^(t) helps identify relevant values of t for extremes and inflections as described for the derivative of R^S^(t).

The effect of any factor possibly modifying the epidemic trend will become apparent first from the changes in the trend of the transmissibility that will modify the epidemic curve.

Considering the curve of symptomatic COViD-19 cases, symptoms are expected to appear seven days after the initial infection^8^; therefore, any detectable effect is expected to appear after the same amount of time. Hospitalizations are expected to occur five days after the symptoms onset^9^. The detection of New SARS-CoV-2 infections is expected to occur 14 days after the infection^10^ due to the delay in detecting and recording the positive cases. Defining t_0_ as the day of introduction of the tiered restrictions, t_0_ + d refers to the start date of the potential effectiveness of the restrictions. The value attributed to d being seven days for the start of symptoms, 12 days for the hospitalizations, and 14 days for the detection of new infections.

## Results

During a period starting in early October, approximately 37-42 days before the expected effectiveness of the restrictions and ending 7-9 days after such time, R(t) has constantly been decreasing with a slope that reached its maximum 24-31 days before the expected effectiveness of the restrictions (Table 1 and Figure 1). This in a consistent way for all the three parameters that were considered: the daily number of new symptomatic cases and new hospitalizations from the ISS database, and the number of new detected SARS-CoV-2 infections from the database of PC.

**Table 1.** Italy - New Cases of COViD-19 by onset of symptoms and New Hospitalizations due to COViD-19. Trend of R*(t) October-November 2020.

| Event | $d^{\S}$ | $R^S(t)$ Decrease Period | | | | | Maximum $R^S(t)$<br>slope decrease | | $R^S(t)=1$ | |
| --- | --- | --- | --- | --- | --- | --- | --- | --- | --- | --- |
| | | Duration | Start | Days<br>before<br>$t_0+d^{\S}$ | End | Days<br>after<br>$t_0+d^{\S}$ | Date | Days<br>before<br>$t_0+d^{\S}$ | Date | Days<br>before<br>$t_0+d$ |
| Symptoms Onset | 7 | 47 | 4Oct20 | 40 | 20Nov20 | 7 | 20Oct20 | 24 | 6Nov20 | 7 |
| Hospitalization | 12 | 44 | 12Oct20 | 37 | 25Nov20 | 7 | 23Oct20 | 26 | 11Nov20 | 7 |
| New SARS-CoV-2<br>infection | 14 | 47 | 13Oct20 | 38 | 29Nov20 | 9 | 27Oct20 | 24 | 14Nov20 | 6 |

<sup>§</sup> d refers to the lag between the onset of containment measures and the start date of their potential effectiveness on the event.

**Figure 1.**
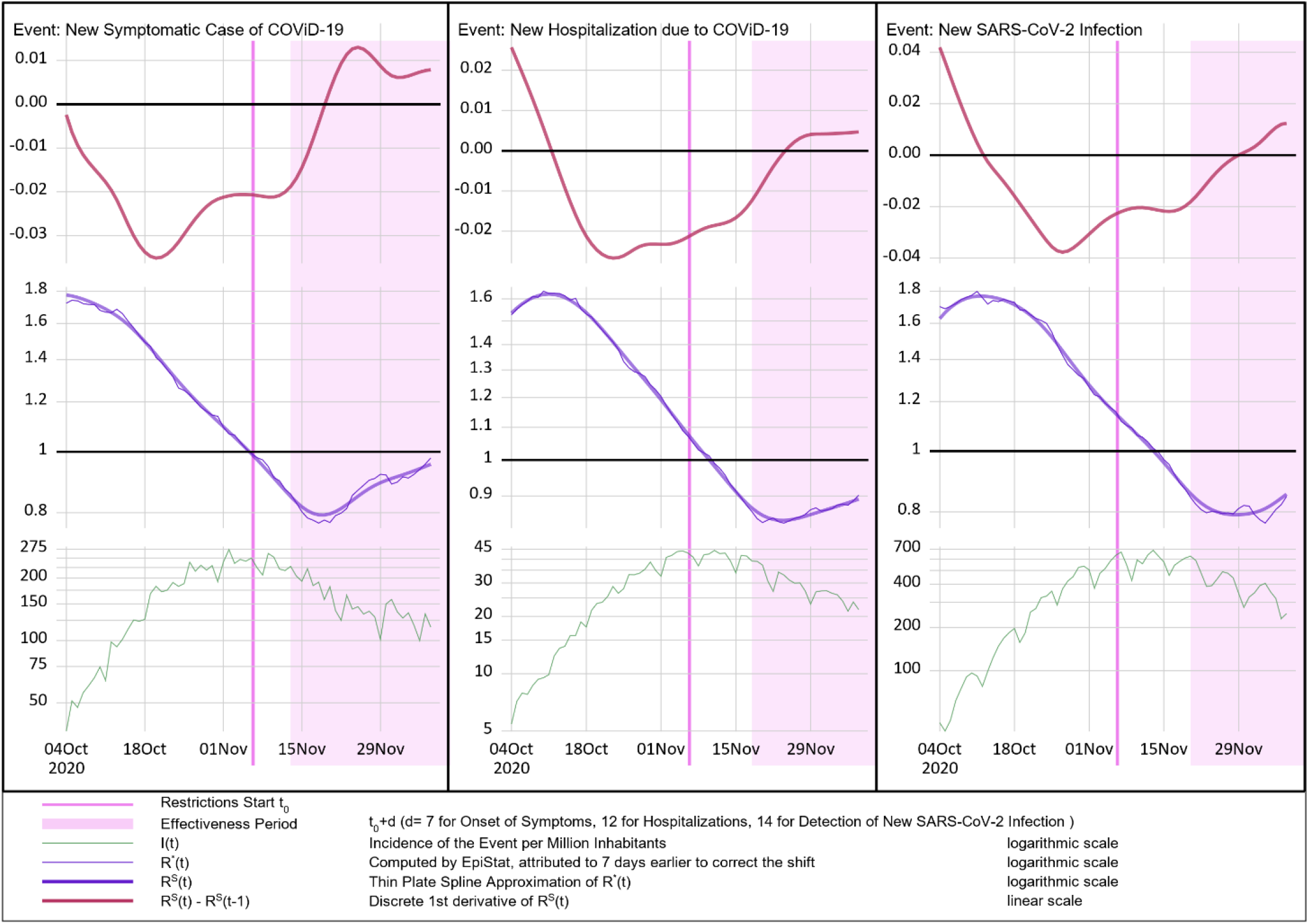
Italy - New Cases of COViD-19 by onset of symptoms, New Hospitalizations due to COViD-19, and New SARS-CoV-2 Infections. Trend of R*(t) October-November 2020 and its 1^st^ derivative.

While irrelevant for this analysis, dealing with the trend of R(t) rather than with any specific value, R^S^(t)=1 was considered to be of special interest for being the time when the epidemic curve I(t) starts decreasing. This evidences that even the outbreak peak was observed before the measures could have an effect. Worsening in the trend of R^S^(t) occurred 7 to 9 days after the start of the period of effectiveness of the measures.

The supportive analyses using G^S^(t) (Table 2 and Figure 2), limited to the pool of all country’s data, using the database of ISS concerning the onset of symptoms and hospitalizations and the database of PC for the report of new SARS-CoV-2 infections, confirm these results.

**Table 2.** Italy - New Cases of COViD-19 by onset of symptoms and New Hospitalizations due to COViD-19. Trend of G^S^(t) October-November 2020.

| Event | $d^{\S}$ | $G^S(t)$ Decrease Period | | | | | Maximum $G^S(t)$<br>slope decrease | | $G^S(t) = 1$ | |
| --- | --- | --- | --- | --- | --- | --- | --- | --- | --- | --- |
| | | Duration | Start | Days<br>before<br>$t_0+d^{\S}$ | End | Days<br>after<br>$t_0+d^{\S}$ | Date | Days<br>before<br>$t_0+d^{\S}$ | Date | Days<br>before<br>$t_0+d$ |
| Symptoms Onset | 7 | 50 | 2Oct20 | 42 | 21Nov20 | 8 | 13Oct20 | 31 | 5Nov20 | 8 |
| Hospitalization | 12 | 49 | 9Oct20 | 40 | 27Nov20 | 9 | 21Oct20 | 28 | 10Nov20 | 8 |
| New SARS-CoV-2<br>infection | 14 | 50 | 16Oct20 | 35 | 5Dec20 | 15 | 22Oct20 | 29 | 11Nov20 | 9 |

<sup>§</sup> d refers to the lag between the onset of containment measures and the start date of their potential effectiveness on the event.

**Figure 2.**
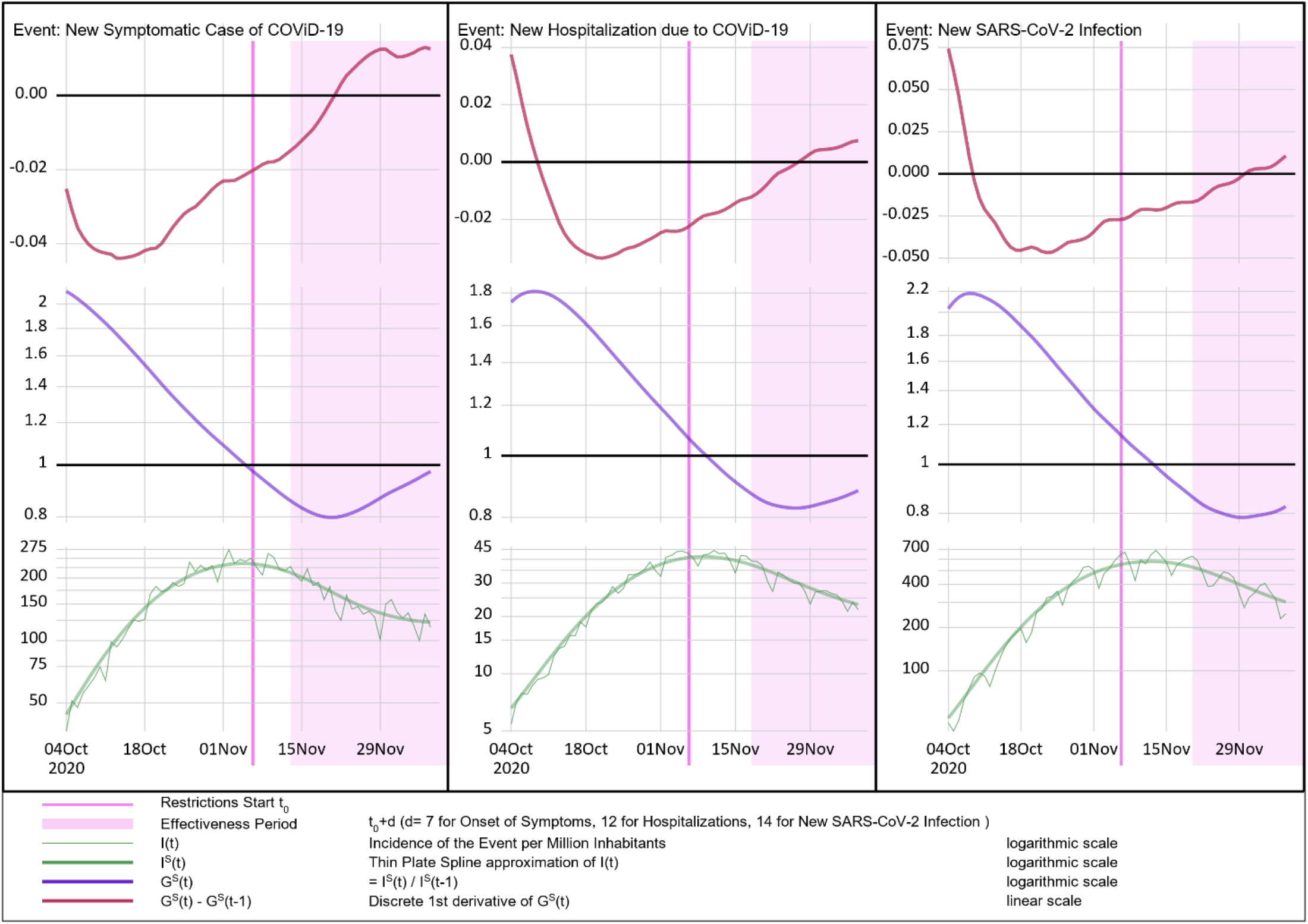
Italy - New Cases of COViD-19 by onset of symptoms and New Hospitalizations due to COViD-19. Trend of G^S^(t) during October-November 2020 and its 1^st^ derivative.

Table 3 and Figure 3 report and display the trend of R*(t) for new SARS-CoV-2 infections detection data provided by PC at regional level for the nine regions with a population exceeding 2 million inhabitants. The outcomes fully match what was obtained at the country level. While this analysis is meant to evaluate the effect of the tiered restrictions as a global intervention, one might consider that the individual tiers to which regions were assigned did not appear to play a relevant role. On 06 November 2020 two regions (Lombardia and Piemonte) moved straight to the strongest (red) tier, two regions (Sicilia and Puglia) moved to the middle tier (orange), while all the other reported regions moved to the mild tier (yellow).

**Table 3.** New SARS-CoV-2 infections in Italy’s regions with more than 2 millions inhabitants. Trend of R^*^(t) October-November 2020.

| | | $R^s(t)$ Decrease Period | | | | | Maximum $R^s(t)$<br>slope decrease | | $R^s(t)=1$ | |
| --- | --- | --- | --- | --- | --- | --- | --- | --- | --- | --- |
| Event | Population<br>(Millions) | Duration | Start | Days<br>before<br>$t_0+d^s$ | End | Days<br>after<br>$t_0+d^s$ | Date | Days<br>before<br>$t_0+d^s$ | Date | Days<br>before<br>$t_0+d^s$ |
| Lombardia<sup>†</sup> | 10,0 | 53 | 10Oct20 | 41 | 2Dec20 | 12 | 28Oct20 | 23 | 11Nov20 | 9 |
| Lazio | 5,7 | 49 | 15Oct20 | 36 | 3Dec20 | 13 | 24Oct20 | 27 | 16Nov20 | 4 |
| Campania | 5,7 | 48 | 15Oct20 | 36 | 2Dec20 | 12 | 2Nov20 | 18 | 11Nov20 | 9 |
| Veneto | 4,9 | 33 | 21Oct20 | 30 | 23Nov20 | 3 | 28Oct20 | 23 | 17Nov20 | 3 |
| Sicilia<sup>‡</sup> | 4,8 | 57 | 11Oct20 | 40 | 7Dec20 | 17 | 16Nov20 | 4 | 19Nov20 | 1 |
| Emilia-Romagna | 4,4 | 40 | 21Oct20 | 30 | 30Nov20 | 10 | 28Oct20 | 23 | 18Nov20 | 2 |
| Piemonte<sup>†</sup> | 4,3 | 54 | 12Oct20 | 39 | 5Dec20 | 15 | 22Oct20 | 29 | 13Nov20 | 7 |
| Puglia<sup>‡</sup> | 3,9 | 59 | 21Oct20 | 30 | 19Dec20 | 29 | 28Oct20 | 23 | 2Dec20 | -12 |
| Toscana | 3,7 | 38 | 19Oct20 | 32 | 26Nov20 | 6 | 28Oct20 | 23 | 11Nov20 | 9 |

<sup>§</sup> d refers to the lag between the onset of containment measures and the start date of their potential effectiveness on the event.
<sup>†</sup> Moved to the strongest containment measures (red) on November 6<sup>th</sup>.
<sup>‡</sup> Moved to the moderate containment measures (orange) on November 6<sup>th</sup>.

**Figure 3.**
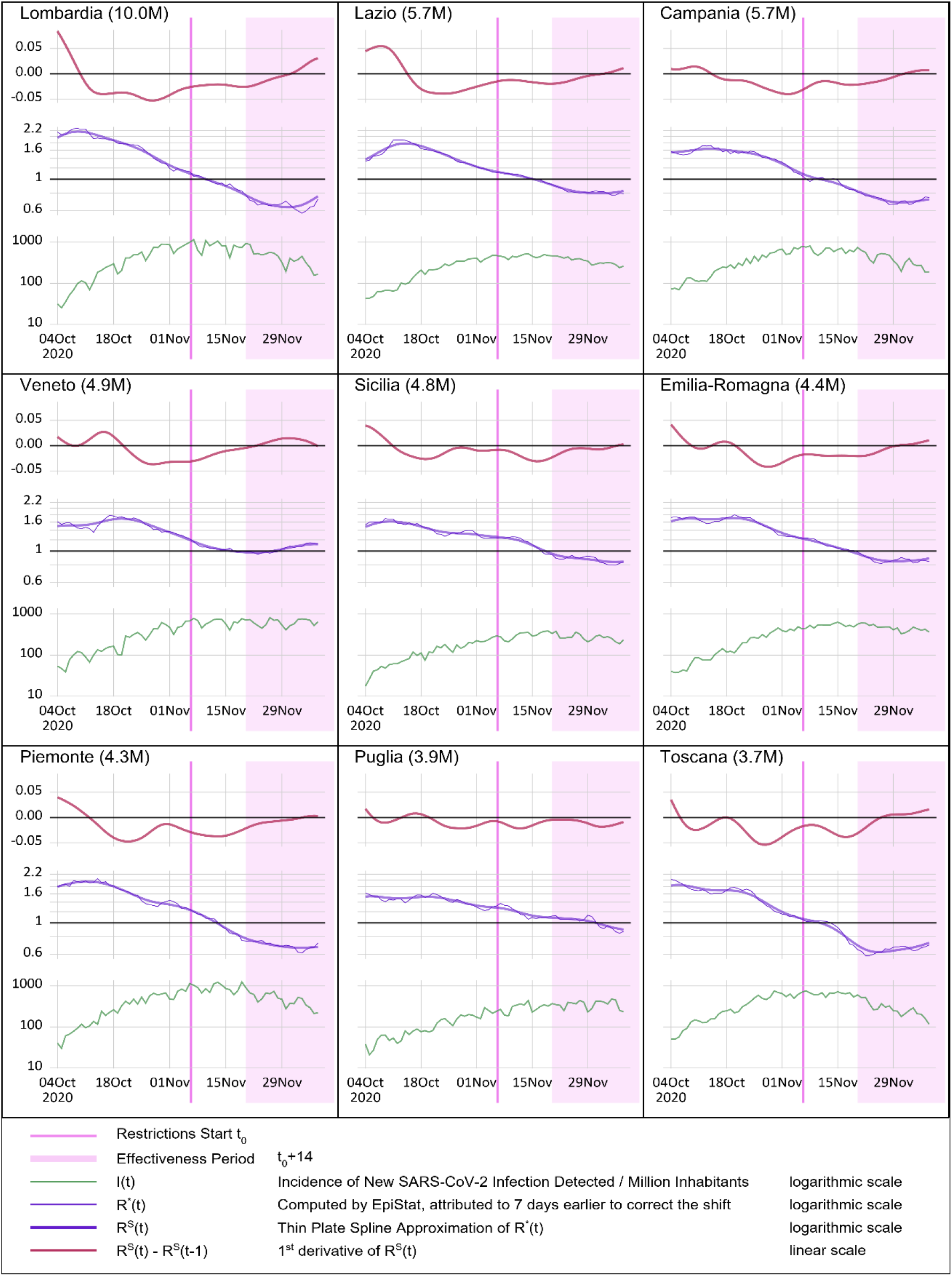
Italy and its most populated tegions - Event: New SARS-CoV-2 infections detection. Trend of R^*^(t) during October-November 2020 and its 1^st^ derivative.

## Discussion

Effective control measures are expected to result in a decreasing reproduction factor’s trend. Changes in the trend direction and slope’s changes are of greatest relevance as they help interpreting changes in the of the epidemics curve that is a mere consequence of these. Positive changes have been observed in the period under study, but this started invariably before introducing the tiered restrictions; moreover, the steepest descending slope was observed before that time. There is no evidence suggesting that the introduction of the countrywide tiered restrictions on 06 November 2021 could have positively modified the reproduction factor’s decreasing trend; therefore, it did not prove to be effective.

The trend of R(t) has started increasing 7 to 9 days after the start of the period of effectiveness of the measures; this does not allow to exclude a negative effect of the measures.

## Limitations

While the need to anticipate by 7 days the R(t) curve estimated by EpiEstim is not described in the literature, it appears to be required to report unit R(t) estimates that are contemporary to the minima and maxima of the epidemic curve to the correct time. R(t) itself can be seen as an approximation of the antilog of the derivative of the log of the incidence I(t), therefore its value in correspondence to the extremes of I(t) must be the unit.

More detailed data at the level of the region and of the province as well as by gender, age and concomitant diseases status would allow a more in-depth analyses, unfortunately these are not made available to the researchers.

There are doubts about the reliability of the data provided by ISS, the Italian Health Institute. While no double check is possible for the data used in this paper, other data, specifically mortality, are reported with gross inconsistencies between and within the institutions: ISS and PC. The size of the differences makes them hard to justify, by different methods of collection, or dates attributions. Estimates derived from three different parameters from two different sources of information have been used to overcome this issue. The smoothing factor of the thin plate spline approximation was chosen arbitrarily. When using alternative values within a reasonable range the results keep qualitatively invariated.

## Conclusions

The Italian data show that the decrease in the trend of R(t) derived from three different parameters from two different sources was independent of the general introduction of tiered restrictions having always preceded it while worsening has been observed one week after the measures became effective. There is no evidence of a positive effect; an unfavorable effect of the tiered containment measures cannot be excluded.

The principle and the nature of the tiered restrictions might require a careful review as they did not seem to have exercised a mitigating influence on the epidemic when they were first introduced nationwide on November 6^th^ 2020. The hint that they might have contributed to boost the epidemics, would suggest performing a much deeper and competent research than what has been done until now. Making detailed data available to independent researchers rather than only to ISS insiders might help in this effort.

## Data Availability

All data are in the public domain, and appropriate reference is made to all sources

https://www.epicentro.iss.it/coronavirus/open-data/covid_19-iss.xlsx

https://github.com/pcm-dpc/COVID-19/tree/master/dati-regioni

